# Multimodal multi-center analysis of electroconvulsive therapy effects: brainwide gray matter increase without functional changes

**DOI:** 10.1101/2022.04.19.22273662

**Authors:** LA van de Mortel, WB Bruin, RM Thomas, C Abbott, M Argyelan, P van Eijndhoven, P Mulders, K Narr, I Tendolkar, JPAJ Verdijk, JA van Waarde, H Bartsch, L Oltedal, GA van Wingen

## Abstract

**Background:** Electroconvulsive therapy (ECT) is an effective treatment for severe depression and induces gray matter (GM) increases in the brain. Small-scale studies suggest that ECT also leads to changes in brain functioning, but findings are inconsistent. In this study, we investigated the influence of ECT on changes in both brain structure and function and their relation to clinical improvement using multicenter neuroimaging data from the Global ECT-MRI Research Collaboration (GEMRIC).

**Methods:** We analyzed T1-weighted structural magnetic resonance imaging (MRI) and functional resting-state MRI data of 88 individuals (49 male) with treatment-resistant depression before and within two weeks after ECT. We performed voxel-based morphometry on the structural data and calculated fractional amplitudes of low-frequency fluctuations, regional homogeneity, degree centrality, functional connectomics, and hippocampus connectivity for the functional data in both unimodal and multimodal analyses. Longitudinal effects in the ECT group were compared to repeated measures of healthy controls (n=27).

**Results:** Wide-spread increases in GM volume were found in patients following ECT. In contrast, no changes in any of the functional measures were observed, and there were no significant differences in structural or functional changes between ECT responders and non-responders. Multimodal analysis revealed that volume increases in the striatum, supplementary motor area and fusiform gyrus were associated with local changes in brain function.

**Conclusion:** These results confirm wide-spread increases in GM volume, but suggest that this is not accompanied by functional changes or associated with clinical response. Instead, focal changes in brain function appear related to individual differences in brain volume increases.

## Introduction

Electroconvulsive therapy (ECT) is the most effective treatment for acute episodes in major depressive disorder (MDD). ECT involves the administration of brief electrical pulses to the brain in anesthetized patients in order to induce generalized seizures. However, ECT is also associated with side-effects such as transient memory loss and other cognitive deficits [1,2], and is therefore typically only used for very severe or treatment-resistant patients. Although the treatment protocol for ECT has drastically changed over the decades and possible side effects have been significantly reduced, further research into ECT is warranted to understand the working mechanism, reduce the stigma that is associated with it, and further optimize the treatment.

Despite ECT being one of the oldest treatments for clinical depression [3], little is known about the biological effects of ECT on the brain and the subsequent alleviations of clinical symptoms. Longitudinal neuroimaging studies using magnetic resonance imaging (MRI) on patients undergoing ECT show an increase in hippocampal gray matter volume [4,5], specifically in the dentate gyrus which is known to be involved in neurogenesis. Many theories on the working mechanism of ECT therefore assume a neuro-, synapto- and/or angiogenic process [6-9], which in turn could lead to structural increases in brain volumes in several regions following ECT such as the temporal cortex and anterior cingulate cortex [10]. However, some studies have shown that these structural increases revert to baseline six months after ECT [11], and recent large-scale studies are inconclusive on whether structural change correlates with symptom improvement [12-14]. These large structural increases with different possible causes therefore seem unable to explain the beneficial effects of ECT.

Besides brain structure, multiple studies have shown an effect of ECT on brain function. When measuring brain function with resting-state fMRI, studies generally use local functional parameters such as the fractional amplitude of low-frequency fluctuations (fALFF), and regional homogeneity (ReHo) or less local parameters such as functional connectivity (FC) and degree centrality (DC). For example, one study reported increased ALFF values in the anterior cingulate cortex and middle frontal gyrus following ECT while decreased values in the precentral gyrus and superior frontal gyrus also have been reported [15-16]. Proximal to these regions, ALFF and DC have shown to increase in the dorsomedial/lateral prefrontal cortex, as well as the bilateral orbitofrontal cortex following ECT, all of which are involved in reward and executive networks [17]. Furthermore, an increase in FC in the right hippocampus to the left superior temporal lobe correlated with symptom improvement [18], as well as an increase in FC from the anterior cingulate cortex to hippocampal, orbitofrontal cortex, and temporal pole regions [15]. However, those findings were typically based on small samples (with N < 30), and/or did not include a control group. The reported changes in brain function may therefore reflect unstable effects due to small sample sizes, or brain changes that are influenced by test-retest effects in healthy controls. An additional limitation of such studies is the focus on one particular brain imaging measure, while the interaction between structural and functional changes may better explain the longer lasting clinical effects of ECT [19]. This multimodal relation, however, has received little attention and thus warrants replication with larger multicenter data.

In this study, we aimed to gain further insight into the neurobiological processes underlying ECT by investigating the relation between structural and functional alterations in patients with depressive episodes using longitudinal structural and functional MRI scans from four treatment sites in The Global ECT-MRI Research Collaboration (GEMRIC). Data from these patients were compared to longitudinal data from healthy controls.

## Methods

### Participants

We initially used longitudinal pre- and post ECT neuroimaging data from four GEMRIC sites including 120 patients. After quality control of the MRI data, we excluded 32 patients that did not meet head motion criteria (rotation/translation < 4mm/degrees, average Framewise Displacement (FD)<0.3 mm and/or subjects having more than 4 minutes of fMRI data with FD<0.25mm) and/or imaging quality criteria (limited signal dropout, no artifacts, satisfactory EPI signal-to-noise ratio). The final sample used in this study thus included 88 patients (49 male, median±IQR of age: 51.5±22.75) diagnosed according to ICD-10 with MDD without psychotic symptoms (n=67), MDD with psychotic symptoms (n=13) or bipolar affective disorder without psychotic symptoms and a current depressive episode (n=9). All patients fulfilled the criteria for moderate to severe depression as measured by the Montgomery-Åsberg Depression Rating Scale (MADRS) ranging from 0-60 (mean score: 34.5 ± 9). Additionally, we used longitudinal structural and functional data from healthy controls (n=27, 11 female) available from one GEMRIC site. These were used to compare longitudinal brain changes between controls and patients after the ECT course. All contributing sites received ethics approval from their local ethics committee or institutional review board. In addition, the centralized mega-analysis was approved by the Regional Ethics Committee South-East in Norway (No. 2018/769).

### Electroconvulsive therapy

For details on site-specific ECT procedures, see [20]. In short, all patients underwent multiple sessions of either right unilateral (RUL, n=53, number (mean±SD) of sessions:10.5±2.4), RUL and bitemporal (n=15, RUL: 6.8±2.6, bitemporal: 5.3±3.9), bitemporal (n=13, 17.6±6.3), RUL and bifrontal (n=1, RUL: 6, bifrontal: 3), or bifrontal only (n=6, 6.8±3.2) stimulations. All post-ECT scans were conducted within two weeks after the last ECT session.

### Imaging

Data was acquired on one Siemens Avanto 1.5T scanner and three 3T scanners (two Siemens Allegra 3T and one General Electric HDx 3T). T1-weighted structural MRI scans were acquired with the following parameters: resolution: 1.0/1.3 × 1.0 × 1.0/1.2 mm3, TR (ms)=2530/2530/7.5/2250, TE=5.16/1.64/3/3.68, T1 (ms) = 1260/1200/450/850, FOV (mm)=256*256*176, flip angle (°)=70/7/8/15. For the resting-state fMRI data, imaging parameters from the four treatment sites were as follows: TR (ms) =2000/2000/2000/1870, TE (ms) =30/29/30/35, flip angle (°)=70/75/*/80,FOV (mm)=240/240/240/240 number of volumes=180/154/150/266,voxel size (mm)=[3.4*3.4*5]/[3.75*3.75*4.55]/[3.75*3.75*3]/[3.5*3.5*3]. For further details regarding the scanning parameters and imaging protocol, see [20].

### Pre-processing

Pre-processing of the structural data was performed using the Computational Anatomy Toolbox [21] for Statistical Parametric Mapping (SPM) software (Wellcome Trust Centre for Neuroimaging, London, UK) in Matlab R2019a [22]. Structural volumes were first skull-stripped and segmented into gray matter (GM), white matter, and cerebrospinal fluid (CSF). With this segmentation, we also calculated each participant’s total intracranial volume (TIV). Each segmented gray matter map was then normalized to the Montreal Neurological Institute (MNI) 1.5 mm template using Diffeomorphic Anatomical Registration using Exponentiated Lie algebra (DARTEL) registration [23] and smoothed with an 8 mm^3^ full-width at half-maximum (FWHM) gaussian kernel to improve the signal-to-noise (SNR) ratio.

Pre-processing of the functional neuroimaging data and the calculation of functional imaging parameters was performed using SPM12 and the Rest Software toolbox in Matlab [24]. First, we removed the first 10 time-points in the functional volumes to achieve steady-state magnetic signal and account for the participant’s situational adaptation. Subsequently, the scans were realigned, coregistered to the structural scan, normalized to Montreal Neurological Institute (MNI) space, resampled to 3 mm isotropic resolution and smoothed with a 6 mm^3^ full-width-at-half-maximum (FWHM) kernel. Smoothing was performed before fALFF/FC and after ReHo/DC calculation to increase the signal-to-noise ratio. Additionally, nuisance covariate regression was performed with Friston’s 24 head-motion parameters, white matter and CSF signal. Lastly, the volumes were bandpass filtered to only include the functional signal at 0.01-0.08 Hz.

### Functional measures

For fALFF, each voxel’s time series was converted to the corresponding fALFF value using the standard calculation procedure [25]. For ReHo, we used the non-smoothed volumes and calculated the Kendall coefficient of concordance (KCC) by voxel-wise comparison of each voxel’s time series to its 27 nearest-neighbors and subsequently smoothed with a 6 mm FWHM kernel.

For DC, we calculated the correlation of each voxel’s time course within a standard MNI gray matter mask with the time course of all other (gray matter) voxels in the brain. Binarized Pearson correlation values above a set threshold of >0.25 were included to calculate a correspondence map for all voxels and smoothed with a 6 mm kernel. Functional connectomics was performed using 116 regions-of-interest (ROI) from the Automatic Anatomical Labeling (AAL) atlas. Each participant’s difference matrix of the connectivity matrices of both time-points was used in the Network-Based Statistics (NBS) toolbox [26] in Matlab.

For functional connectivity, the mean time-series was extracted from the cluster that showed the largest structural change in the VBM analysis, defined as a sphere with a 12 mm radius around the highest statistical peak. A Pearson correlation coefficient between the region’s time-series and each voxel’s time course outside the seed region was calculated. Fisher’s r-to-z transformation was applied to the correlation map.

### Statistical analysis

Clinical MADRS score pre- and post ECT and demographic variables between groups were compared using a Chi-Squared Test and Wilcoxon Signed-Rank Sum test in R (R Core Team, 2021) with an α of .05.

For unimodal statistical analysis of structural and functional brain changes following ECT, we used factorial ANOVAs in SPM12 with the factors time (before ECT, 1-2 weeks after ECT) and group (responders, non-responders, healthy controls) while accounting for the covariates age, sex, treatment site, electrode location (right unilateral, bitemporal, or bifrontal), number of ECT sessions, and additionally TIV for VBM. We employed whole-brain family wise error (FWE) rate correction for multiple voxel-wise comparisons using threshold-free cluster enhancement (TFCE) at a significance threshold of 0.05. Two planned Helmert contrasts were used to assess 1) differences over time between both ECT groups and healthy controls and 2) differences over time within the ECT group between clinical responders vs. non-responders.

Correlation analysis between brain changes in structure/function and clinical improvement were performed with multiple linear regression analyses in SPM using the difference map post-pre for the VBM and functional data, and relevant clinical variables (MADRS score reductions, controlling for the same variables as described above).

For functional connectomics, we conducted a one-way ANOVA in the NBS toolbox to test for any group-specific differences in network FC over time (T-threshold: 3.1, 5000 permutations, α=0.05, results calculated based on network extent).

### Multimodal neuroimaging data analysis

Correlations between structural and functional brain changes were assessed with linear regression models using the VoxelStats package in Matlab [27]. We assessed relations between structural and functional changes for each functional measure separately with the following linear model:

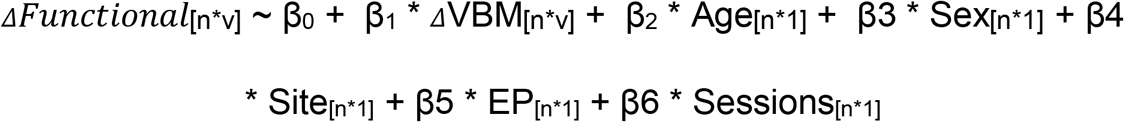

Where n is the total number of patients (88), v is the number of voxels in the imaging modality, EP is electrode placement and sessions is the number of sessions. Whole brain statistical inference was performed on the basis of Random Field Theory (RFT) on the cluster-level using a cluster-defining threshold of p < 0.001 (p < 0.05, FWE-corrected).

## Results

### Group demographics

The patient group had a significantly lower median age (51.5, IQR=22.8) than the healthy control group (61, IQR=12, p=0.002).There were no significant differences in sex between the two groups (X^2^ (2) =.11, p = 0.915). The Wilcoxon Signed-Rank Test confirmed that MADRS scores were significantly lower after (median=14.6, IQR=20.5) than before ECT (median=34,IQR=11.6)), p=<0.001. For full group demographics, see table 1.

**Table 1:**
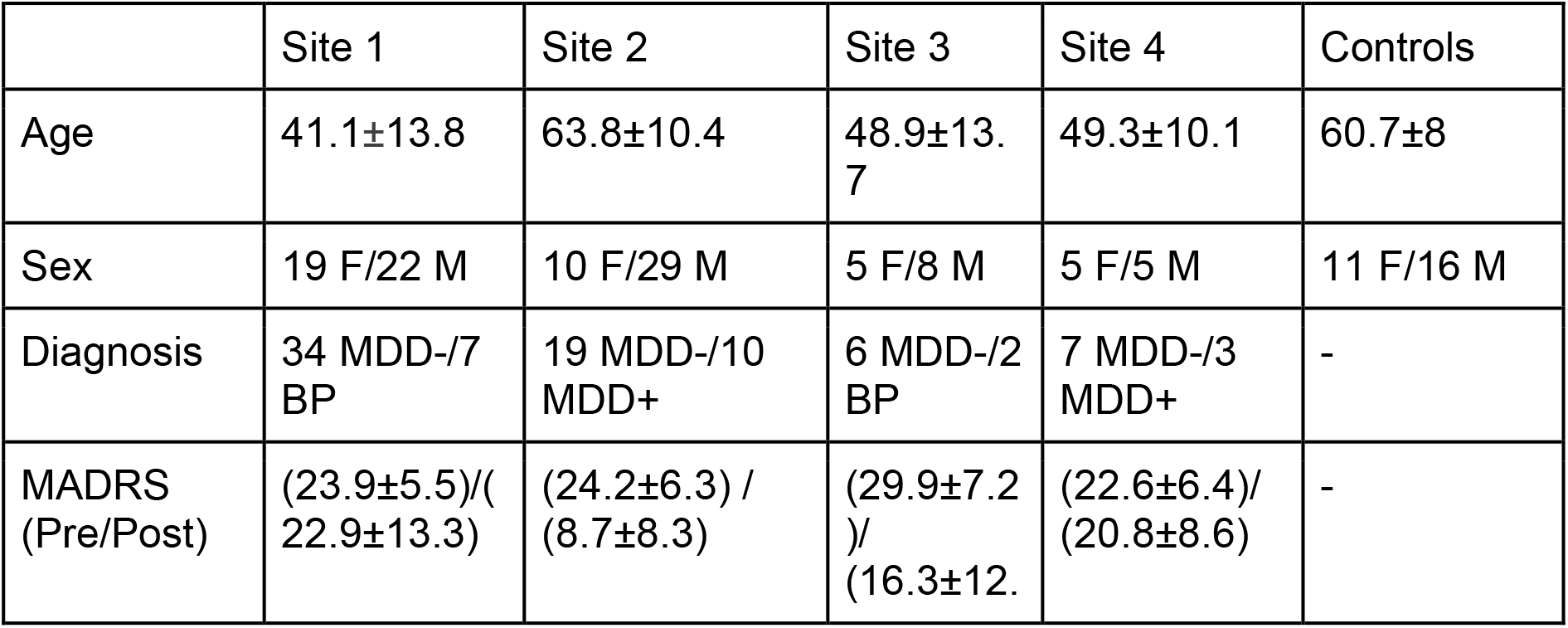
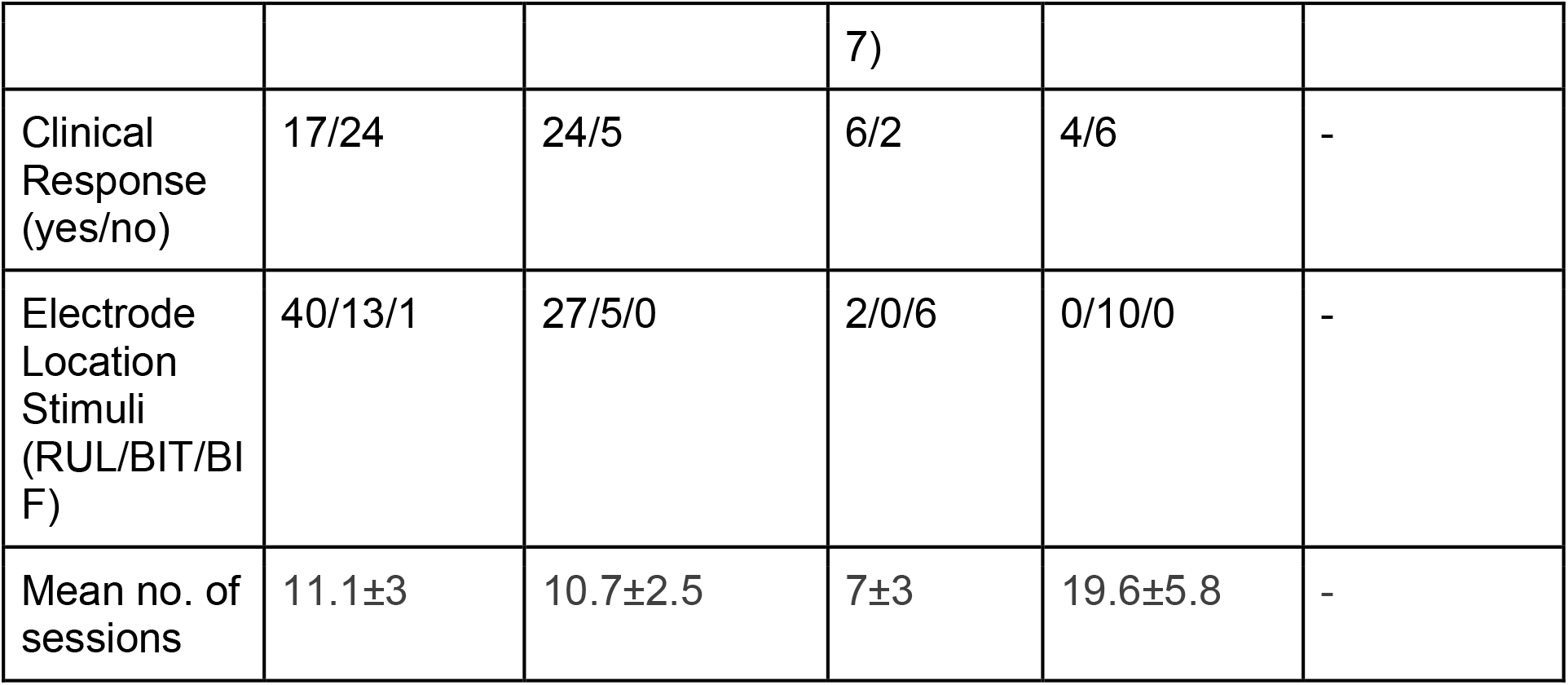
Group demographics of patients (n=88) and healthy controls (n=27). Table includes Age, Sex, Diagnosis (MDD with/without psychotic symptoms (MDD+/-), bipolar disorder (BP)), MADRS scores, Clinical Response (>50% reduction in clinical scores), electrode location stimulation, and mean number of ECT sessions.

|  | Site 1 | Site 2 | Site 3 | Site 4 | Controls |
| --- | --- | --- | --- | --- | --- |
| Age | 41.1±13.8 | 63.8±10.4 | 48.9±13.7 | 49.3±10.1 | 60.7±8 |
| Sex | 19 F/22 M | 10 F/29 M | 5 F/8 M | 5 F/5 M | 11 F/16 M |
| Diagnosis | 34 MDD-/7 BP | 19 MDD-/10 MDD+ | 6 MDD-/2 BP | 7 MDD-/3 MDD+ | - |
| MADRS (Pre/Post) | (23.9±5.5)/(22.9±13.3) | (24.2±6.3) / (8.7±8.3) | (29.9±7.2) / (16.3±12. | (22.6±6.4)/(20.8±8.6) | - |
|  |  |  | 7) |  |  |
| Clinical Response (yes/no) | 17/24 | 24/5 | 6/2 | 4/6 | - |
| Electrode Location Stimuli (RUL/BIT/BIF) | 40/13/1 | 27/5/0 | 2/0/6 | 0/10/0 | - |
| Mean no. of sessions | 11.1±3 | 10.7±2.5 | 7±3 | 19.6±5.8 | - |

### Unimodal analysis

The group x time interaction VBM analysis revealed wide-spread increases in GM volume in patients following ECT compared to controls. One large cluster of 310846 voxels with the peak in the right parahippocampal gyrus extended bilaterally to the thalamus, left parahippocampal gyrus, midfrontal areas, superior parietal lobe, and occipital and cerebellar areas. Additional smaller clusters were widely distributed in the brain (see Figure 1 and Table 2).The comparison between clinical responders and non-responders revealed no significant differences in gray matter increase. Remarkably, the group x time interaction revealed no significant differences in functional changes between patients and HC for fALFF, ReHo, DC, and functional connectomics. FC changes following ECT were assessed by placing the seed for the FC analysis in the highest statistical peak in the right (para)hippocampus. The group x time interaction showed no significant changes in functional connectivity between patients and controls. And the group x time interaction between responders and non-responders revealed no significant differences for any of the functional measures. We further investigated whether these null findings were related to differences across research sites or disease diagnosis. These additional analyses, excluding patients with psychotic symptoms and bipolar affective disorder yielded similar (non-significant) results. And there were no significant differences in functional changes between treatment sites as assessed by an additional factorial ANOVA with site as factor. Furthermore, to investigate whether there were any observable longitudinal brain changes in patients at all that were common among responders and non-responders, we compared the functional measures before and after treatment across both patient groups. However, none of the results from this contrast on any of the functional measures in the patient group survived statistical correction for multiple comparisons.

**Figure 1.**
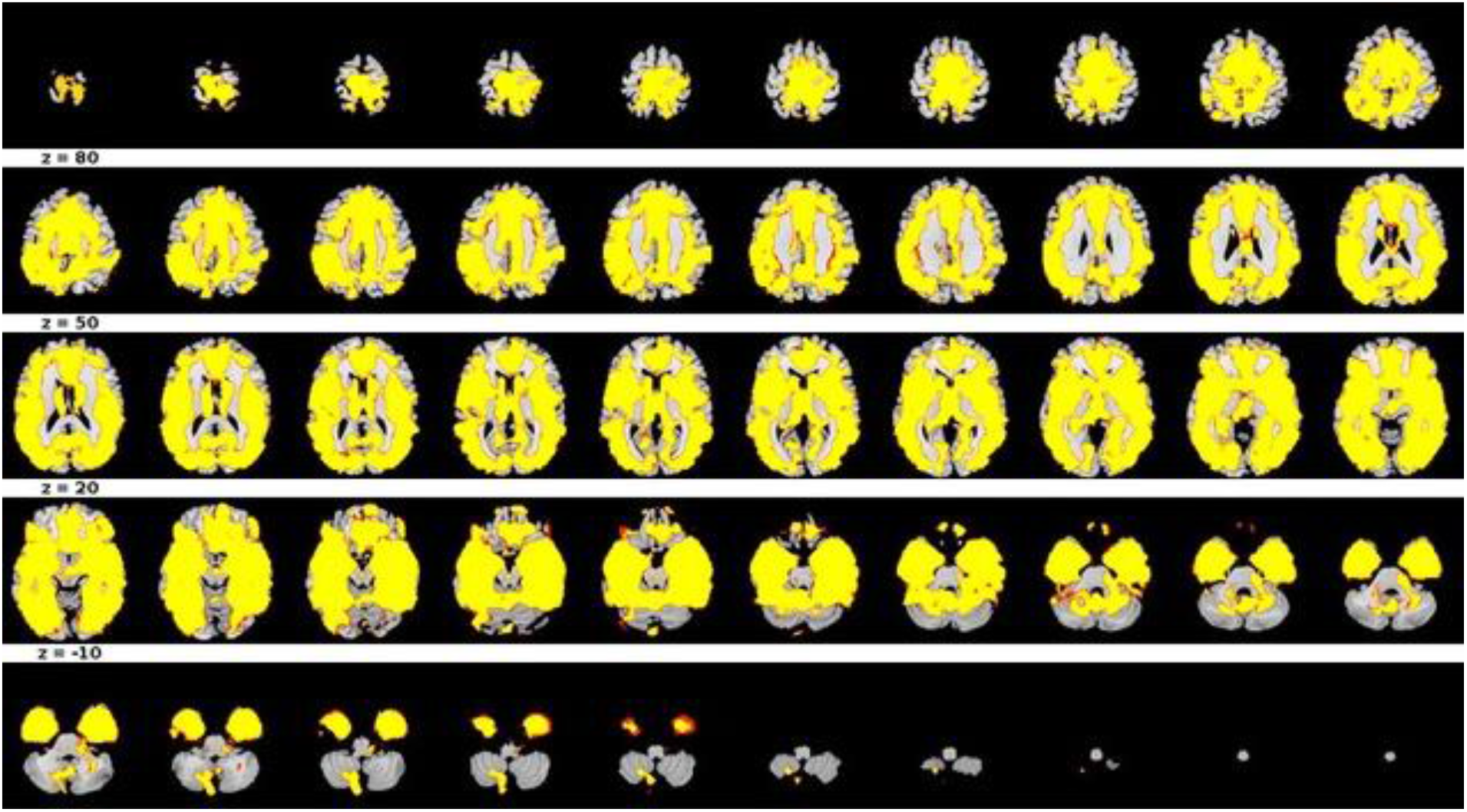
Structural gray matter increases in the brain of patients following ECT compared to longitudinal data of healthy controls.

**Table 2:** Gray matter increases over time in patients receiving ECT compared to healthy controls. Table displays the specific brain region based on the AAL atlas, cluster extent (number of voxels with >5 reported in the table), FWE-corrected p-value, TFCE value, statistical Z-value and MNI coordinates.

|  |  |  |  |  | MNI coordinates (mm) |  |  |
| --- | --- | --- | --- | --- | --- | --- | --- |
| Brain Region | Extent | p (FWE-corr) | TFCE | Z-value | x | y | z |
| ParaHippocampal_R | 310846 | <0.001 | 23347.58 | 3.35 | 28 | 2 | -28 |
| Hippocampus_R |  | <0.001 | 22644.34 | 3.35 | 21 | -6 | -21 |
| Hippocampus_R |  | <0.001 | 21679.49 | 3.35 | 32 | -6 | -18 |
| Cerebellum_10_L | 27 | 0.003 | 840.7 | 3.04 | -12 | -28 | -45 |
| ParaHippoca<br>mpal_L | 20 | 0.004 | 810.07 | 2.95 | -8 | -22 | -21 |
| Parietal_Inf_R | 7 | 0.007 | 253.99 | 2.23 | 57 | -57 | 48 |
| Frontal_Mid_L | 7 | 0.007 | 252.32 | 2.27 | -40 | 48 | 32 |
| Cerebelum_C<br>rus2_R | 251 | 0.007 | 245.18 | 2.37 | 0 | -93 | -24 |
| Cerebelum_C<br>rus2_L |  | 0.008 | 223.03 | 2.15 | -2 | -86 | -21 |
| Cerebelum_C<br>rus2_L |  | 0.009 | 208.38 | 2.16 | -10 | -94 | -26 |

Additionally, to explore whether the level of clinical improvement was related to changes in brain structure and function, we performed unimodal regression analyses between post-pre MADRS scores and post-pre ECT brain difference maps with age, sex, research site, electrode location, and number of sessions as a covariate. These analyses did not reveal any significant relation with clinical improvement for any of the structural and functional measures.

### Multimodal analysis

To assess the correlation between the structural change and functional changes in every voxel independently, we performed additional multimodal analyses. This analysis showed small clusters of significant correlations between changes in gray matter volume and various functional parameters.

First, the gray matter increase in the striatum (caudate nucleus and nucleus accumbens) showed relations with a reduction in DC in this area. Furthermore, the gray matter change in a small cluster in the left fusiform gyrus was related to an increase in fALFF in the same area. Both the left and right supplementary motor area (SMA) showed similar relations between the gray matter increase and fALFF increase (see figure 2).

**Figure 2:**
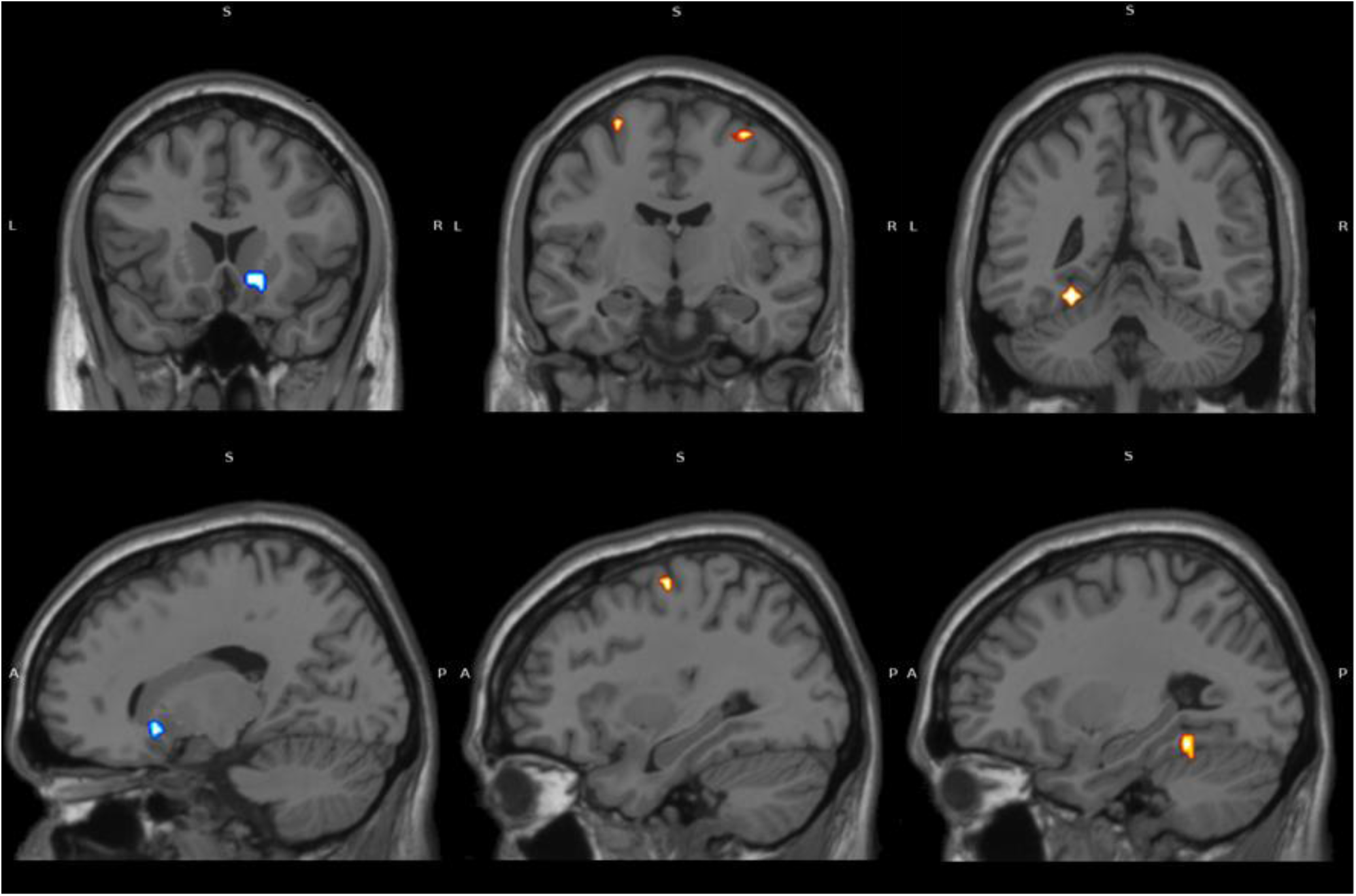
Significant relations between gray matter volume increase and DC decrease in the right ventral striatum (blue,left) and ALFF increase in the bilateral SMA (red, middle) and left fusiform gyrus (red,right)

After extracting the mean changes in structural and functional values of these clusters, we found no significant correlation between gray-matter related changes in fALFF and DC functional values with symptom improvement.

## Discussion

In this longitudinal multicenter study, we investigated the influence of ECT on brain structure and function in severely depressed patients. The structural VBM analysis showed wide-spread increases in GM volume with the peak in the right hippocampal area. Strikingly, we did not observe a consistent change in brain function as measured with resting-state fALFF, ReHo, DC, functional connectomics or hippocampal connectivity. Despite the lack of consistent functional changes, we did find a correlation between the volume increase and distinct changes in local activity in the striatum, fusiform gyrus, and bilateral SMA.

### Structural brain changes

Many studies over the past years have attempted to get better insight into the working mechanism of ECT for depressive symptoms on the structural, functional, and clinical level. Both animal- and human studies point to an effect of ECT on structural gray matter increases in many regions [8, 28-29], but other studies have not always been able to find relations between this structural increase of brain volumes and clinical improvement [5,12,14,30]. In our study, increase in regional brain volume was not confined to the hippocampus and/or medial temporal lobe, as was suggested by the initial studies [4,5,10]. Similar to a recent anatomical parcellation study from GEMRIC [14], the structural increase in patients receiving ECT was widely distributed across the brain. The fact that the highest statistical peak was found in the right hippocampal area could be attributed to the electrode placement and coinciding electric field strength [31-32], presumably due to the majority of the patient sample used in this study receiving RUL stimulation. Again, in line with the previous report from GEMRIC, the large-scale volume increase was not related to clinical response, as indicated by both the factorial ANOVA and multiple regression analysis. While this is in line with aforementioned studies failing to find relations between structural volume increase and symptom improvement, it remains puzzling how these robust structural brain changes seem to have no effect on the depressive mood severity scores. Therefore, our goal was to examine the effect of functional brain changes as well.

### Functional brain changes

Perhaps even more puzzling is the fact that functional measures of local and global synchronization seem largely unaffected by large-scale structural brain volume increases, since no changes in any functional parameter were observed in our patients following ECT, even after controlling for treatment response, treatment site difference, electrode placement, and the number of ECT sessions in the course. In other words, ECT does not seem to cause any consistent changes in the brain on the local level (as measured by fALFF and ReHo), nor on the global network-like level (as measured by DC, and seed-based/network-based FC). This is not in line with previous (small-scale) fMRI studies reporting functional changes in frontal areas and a normalization of hippocampal connectivity following ECT [15,17,18,33]. It should be noted that some of the aforementioned studies lacked a control group in their analyses. This makes it uncertain whether these results were due to clinical effects or test-retest effects commonly seen in longitudinal resting-state fMRI studies [34-36]. Also, these aforementioned studies may have reported findings that generalize poorly due to their low sample size [37]. Importantly, we adopted a more stringent statistical correction and corrected for multiple voxel-wise comparisons across the whole brain, while other studies typically restricted their analysis to particular ROIs. We chose our approach because of the previously reported global effects of ECT on brain structure [14], which we expected to be related to brain-wide changes in brain function. While we consider the stringent correction for multiple comparisons a strength of our study, this approach does not exclude the possibility that smaller effects exist that do not withstand whole-brain correction.

### Multimodal functional analysis

While our unimodal analyses did not detect consistent changes in brain function, our multimodal analysis did show a relation between gray matter increases and striatal DC reductions, and an association between gray matter increases and increases in ALFF in the bilateral SMA and left fusiform gyrus. While the average change in ALFF and DC in these regions did not correlate with symptom improvement, these regions have shown relevance for depressive disorders. The striatum (consisting of the caudate nucleus, putamen, and nucleus accumbens) is involved in reward processing, and aberrant function of the reward circuitry is considered a trait of depressive disorders [38], presumably related to altered striatal function [39-40]. This could also explain why the ventral striatum is an effective target for neurosurgical treatment of depressive disorders [41].

While less is known about the role of the SMA and fusiform gyrus in depressive disorders, a reduction of SMA volume has previously been reported [42], as well as activity normalization of the SMA following pharmacological treatment [43] which could be related to the psychomotor retardation often seen in depressed patients [44]. Similarly, FC of the fusiform gyrus with sensorimotor areas has shown to be reduced in major depressive disorder [4-46] and fusiform gyrus structure could be related to emotional behavior in depression [47].

Thus, while ECT showed no consistent changes in brain function, these analyses suggest that the changes in brain function are dependent on the extent of gray matter increases.

### Limitations

The null results of this study have to be considered in light of certain limitations. One important aspect is the composition of our sample. The healthy control sample was smaller (n=27) and significantly older, and we corrected for variation in age by including it as covariate in the analyses. Although this may have masked more subtle effects, this did not preclude the detection of robust brain-wide changes in GM. Additionally, the patient sample used in this study consisted of multiple diagnoses (MDD with and without psychotic symptoms, bipolar disorder), which could have limited the interpretability of our findings and also decreased our statistical power. Also, the neuroimaging data came from different scanners and treatment sites. While all patients were scanned within two weeks after the last ECT-session, there still will have been some variability between time-points. However, we did not find evidence that either site or diagnosis affected our results, indicating that these results were not due to differences in scanner type, sample characteristics, and time-point after the last ECT session.

A more plausible explanation for our null findings could be that resting-state fMRI is not very sensitive to detect functional effects of ECT. Although the resting-state data used in this study was not optimal and limited by the relatively minimal acquisition time (6 to 8 minutes), and the fact that the effects of ECT on resting-state activity could occur during the entire ECT-course instead of within two weeks after the last session, it may be possible that BOLD-signal fluctuations are just not always sensitive to ECT effects on brain function. Rather, ECT effects could more accurately be studied in brain activity measures such as specific EEG frequency bands [48-49], which is also more analogous to the ictal mechanism behind ECT. The biological processes occurring during and after ECT may also involve other processes such as molecular neurotransmission effects, since alterations in serotonergic 5-HT1A and 5-HT2A receptor binding and transmission following ECT have been reported, both in the hippocampus and frontal affective areas (50-51). As a result, resting-state fMRI data may not accurately reflect the therapeutic effects of ECT. While this multi-modal study aimed to find relations between structural and functional changes in the brain following ECT, future multimodal studies may uncover the relation between brain activity as measured by EEG and fMRI in patients receiving ECT.

Altogether, the group-level null findings in this study are unable to explain the working mechanisms of ECT on a structural or functional level, since structural brain changes seem largely unrelated to changes in brain function and clinical scores. In fact, brain activity as measured with BOLD-signal fluctuations seems almost entirely independent of underlying gray matter structure or subjective mood, since our results show that both brain structure and depressive scores can change drastically while the BOLD-signal during resting-state fMRI acquisition does not change. While we did find an association between structural and functional changes, indicating that functional changes are dependent on ECT-related changes in brain structure, the detected clusters were small and did not correlate with clinical improvement. In conclusion, this multicenter study with a relatively large sample confirmed that ECT leads to wide-spread increases in gray matter volume. However, this was not accompanied by consistent changes in various measures of brain function, and not related to clinical improvement. Our multimodal analysis did reveal an association between the increase in brain volume and changes in regional activity measures, indicating that structural brain changes have only minimal functional consequences on the brain-systems level.

## Data Availability

All data produced in the present work are contained in the manuscript

## Acknowledgements

This study was funded by the Western Norway Regional Health Authority (Grant Numbers 911986 and 912238) and the National Institute for Mental Health (NIMH, USA, Grant Numbers MH125126, MH119616, MH092301 and MH110008). The authors declare no competing financial interests or any other conflict of interest.

## References

1. Tørring, N., Sanghani, S. N., Petrides, G., Kellner, C. H., & Østergaard, S. D. (2017). The mortality rate of electroconvulsive therapy: a systematic review and pooled analysis. Acta Psychiatrica Scandinavica, 135(5), 388–397. https://doi.org/10.1111/acps.12721

2. Hermida, A. P., Glass, O. M., Shafi, H., & McDonald, W. M. (2018). Electroconvulsive Therapy in Depression. Psychiatric Clinics of North America, 41(3), 341–353. https://doi.org/10.1016/j.psc.2018.04.001

3. Gazdag, G., & Ungvari, G. S. (2019). Electroconvulsive therapy: 80 years old and still going strong. World Journal of Psychiatry, 9(1), 1–6. https://doi.org/10.5498/wjp.v9.i1.1

4. Nordanskog, P., Dahlstrand, U., Larsson, M. R., Larsson, E.-M., Knutsson, L., & Johanson, A. (2010). Increase in Hippocampal Volume After Electroconvulsive Therapy in Patients With Depression. The Journal of ECT, 26(1), 62–67. https://doi.org/10.1097/yct.0b013e3181a95da8

5. Joshi, S. H., Espinoza, R. T., Pirnia, T., Shi, J., Wang, Y., Ayers, B., Leaver, A., Woods, R. P., & Narr, K. L. (2016). Structural Plasticity of the Hippocampus and Amygdala Induced by Electroconvulsive Therapy in Major Depression. Biological Psychiatry, 79(4), 282–292. https://doi.org/10.1016/j.biopsych.2015.02.029

6. Bouckaert, F., Sienaert, P., Obbels, J., Dols, A., Vandenbulcke, M., Stek, M., & Bolwig, T. (2014). ECT: its brain enabling effects: a review of electroconvulsive therapy-induced structural brain plasticity. The Journal of ECT, 30(2), 143–151. https://doi.org/10.1097/YCT.0000000000000129

7. Rotheneichner, P., Lange, S., O’Sullivan, A., Marschallinger, J., Zaunmair, P., Geretsegger, C., Aigner, L., & Couillard-Despres, S. (2014). Hippocampal Neurogenesis and Antidepressive Therapy: Shocking Relations. Neural Plasticity, 2014. https://doi.org/10.1155/2014/723915

8. Nuninga, J. O., Mandl, R. C. W., Boks, M. P., Bakker, S., Somers, M., Heringa, S. M., Nieuwdorp, W., Hoogduin, H., Kahn, R. S., Luijten, P., & Sommer, I. E. C. (2020). Volume increase in the dentate gyrus after electroconvulsive therapy in depressed patients as measured with 7T. Molecular Psychiatry, 25(7), 1559–1568. https://doi.org/10.1038/s41380-019-0392-6

9. Van Den Bossche, M. J. A., Emsell, L., Dols, A., Vansteelandt, K., De Winter, F.-L., Van den Stock, J., Sienaert, P., Stek, M. L., Bouckaert, F., & Vandenbulcke, M. (2019). Hippocampal volume change following ECT is mediated by rs699947 in the promotor region of VEGF. Translational Psychiatry, 9(9). https://doi.org/10.1038/s41398-019-0530-6

10. Ota, M., Noda, T., Sato, N., Okazaki, M., Ishikawa, M., Hattori, K., Hori, H., Sasayama, D., Teraishi, T., Sone, D., & Kunugi, H. (2015). Effect of electroconvulsive therapy on gray matter volume in major depressive disorder. Journal of affective disorders, 186, 186–191. https://doi.org/10.1016/j.jad.2015.06.051

11. Nordanskog, P., Larsson, M. R., Larsson, E.-M.., & Johanson, A. (2013). Hippocampal volume in relation to clinical and cognitive outcome after electroconvulsive therapy in depression. Acta Psychiatrica Scandinavica, 129(4), 303–311. https://doi.org/10.1111/acps.12150

12. Oltedal, L., Narr, K. L., Abbott, C., Anand, A., Argyelan, M., Bartsch, H., Dannlowski, U., Dols, A., van Eijndhoven, P., Emsell, L., Erchinger, V. J., Espinoza, R., Hahn, T., Hanson, L. G., Hellemann, G., Jorgensen, M. B., Kessler, U., Oudega, M. L., Paulson, O. B., & Redlich, R. (2018). Volume of the Human Hippocampus and Clinical Response Following Electroconvulsive Therapy. Biological Psychiatry, 84(8), 574–581. https://doi.org/10.1016/j.biopsych.2018.05.017

13. Mulders, P., Llera, A., Beckmann, C. F., Vandenbulcke, M., Stek, M., Sienaert, P., Redlich, R., Petrides, G., Oudega, M. L., Oltedal, L., Oedegaard, K. J., Narr, K. L., Magnusson, P. O., Kessler, U., Jorgensen, A., Espinoza, R., Enneking, V., Emsell, L., Dols, A., Dannlowski, U., … Tendolkar, I. (2020). Structural changes induced by electroconvulsive therapy are associated with clinical outcome. Brain stimulation, 13(3), 696–704. https://doi.org/10.1016/j.brs.2020.02.020

14. Ousdal, O. T., Argyelan, M., Narr, K. L., Abbott, C., Wade, B., Vandenbulcke, M., Urretavizcaya, M., Tendolkar, I., Takamiya, A., Stek, M. L., Soriano-Mas, C., Redlich, R., Paulson, O. B., Oudega, M. L., Opel, N., Nordanskog, P., Kishimoto, T., Kampe, R., Jorgensen, A., & Hanson, L. G. (2020). Brain Changes Induced by Electroconvulsive Therapy Are Broadly Distributed. Biological Psychiatry, 87(5), 451– 461. https://doi.org/10.1016/j.biopsych.2019.07.010

15. Liu, Y., Du, L., Li, Y., Liu, H., Zhao, W., Liu, D., Zeng, J., Li, X., Fu, Y., Qiu, H., Li, X., Qiu, T., Hu, H., Meng, H., & Luo, Q. (2015). Antidepressant Effects of Electroconvulsive Therapy Correlate With Subgenual Anterior Cingulate Activity and Connectivity in Depression. Medicine, 94(45), e2033. https://doi.org/10.1097/md.0000000000002033

16. Kong, X. M., Xu, S. X., Sun, Y., Wang, K. Y., Wang, C., Zhang, J., Xia, J. X., Zhang, L., Tan, B. J., & Xie, X. H. (2017). Electroconvulsive therapy changes the regional resting state function measured by regional homogeneity (ReHo) and amplitude of low frequency fluctuations (ALFF) in elderly major depressive disorder patients: An exploratory study. Psychiatry research. Neuroimaging, 264, 13–21. https://doi.org/10.1016/j.pscychresns.2017.04.001

17. Zhang, T., He, K., Bai, T., Lv, H., Xie, X., Nie, J., Xie, W., Zhu, C., Wang, K., & Tian, Y. (2021). Altered neural activity in the reward-related circuit and executive control network associated with amelioration of anhedonia in major depressive disorder by electroconvulsive therapy. Progress in Neuro-Psychopharmacology and Biological Psychiatry, 109(264), 110193. https://doi.org/10.1016/j.pnpbp.2020.110193

18. Abbott, C. C., Jones, T., Lemke, N. T., Gallegos, P., McClintock, S. M., Mayer, A. R., Bustillo, J., & Calhoun, V. D. (2014). Hippocampal structural and functional changes associated with electroconvulsive therapy response. Translational Psychiatry, 4(11), e483. https://doi.org/10.1038/tp.2014.124

19. Qi, S., Abbott, C. C., Narr, K. L., Jiang, R., Upston, J., McClintock, S. M., Espinoza, R., Jones, T., Zhi, D., Sun, H., Yang, X., Sui, J., & Calhoun, V. D. (2020). Electroconvulsive therapy treatment responsive multimodal brain networks. Human brain mapping, 41(7), 1775–1785. https://doi.org/10.1002/hbm.24910

20. Oltedal, L., Bartsch, H., Sørhaug, O. J. E., Kessler, U., Abbott, C., Dols, A., Stek, M. L., Ersland, L., Emsell, L., van Eijndhoven, P., Argyelan, M., Tendolkar, I., Nordanskog, P., Hamilton, P., Jorgensen, M. B., Sommer, I. E., Heringa, S. M., Draganski, B., Redlich, R., & Dannlowski, U. (2017). The Global ECT-MRI Research Collaboration (GEMRIC): Establishing a multi-site investigation of the neural mechanisms underlying response to electroconvulsive therapy. NeuroImage: Clinical, 14(14), 422–432. https://doi.org/10.1016/j.nicl.2017.02.009

21. Cat12 version 12.7: http://www.neuro.uni-jena.de/cat/

22. MATLAB and Statistics Toolbox Release 2019a, The MathWorks, Inc., Natick, MA, US

23. Ashburner, J. (2007). A fast diffeomorphic image registration algorithm. NeuroImage, 38(1), 95–113. https://doi.org/10.1016/j.neuroimage.2007.07.007

24. Song, X.-W., Dong, Z.-Y., Long, X.-Y., Li, S.-F., Zuo, X.-N., Zhu, C.-Z., He, Y., Yan, C.-G., & Zang, Y.-F. (2011). REST: A Toolkit for Resting-State Functional Magnetic Resonance Imaging Data Processing. PLoS ONE, 6(9), e25031. https://doi.org/10.1371/journal.pone.0025031

25. Zou, Q.-H., Zhu, C.-Z., Yang, Y., Zuo, X.-N., Long, X.-Y., Cao, Q.-J., Wang, Y.-F., & Zang, Y.-F. (2008). An improved approach to detection of amplitude of low-frequency fluctuation (ALFF) for resting-state fMRI: Fractional ALFF. Journal of Neuroscience Methods, 172(1), 137–141. https://doi.org/10.1016/j.jneumeth.2008.04.012

26. Zalesky, A., Fornito, A., & Bullmore, E. T. (2010). Network-based statistic: identifying differences in brain networks. NeuroImage, 53(4), 1197–1207. https://doi.org/10.1016/j.neuroimage.2010.06.041

27. Mathotaarachchi, S., Wang, S., Shin, M., Pascoal, T. A., Benedet, A. L., Kang, M. S., … & Rosa-Neto, P. (2016). VoxelStats: a MATLAB package for multi-modal voxel-wise brain image analysis. Frontiers in neuroinformatics, 10, 20.

28. Pirnia, T., Joshi, S. H., Leaver, A. M., Vasavada, M., Njau, S., Woods, R. P., Espinoza, R., & Narr, K. L. (2016). Electroconvulsive therapy and structural neuroplasticity in neocortical, limbic and paralimbic cortex. Translational Psychiatry, 6(6), e832. https://doi.org/10.1038/tp.2016.102

29. Levy, M. J. F., Boulle, F., Steinbusch, H. W., van den Hove, D. L. A., Kenis, G., & Lanfumey, L. (2018). Neurotrophic factors and neuroplasticity pathways in the pathophysiology and treatment of depression. Psychopharmacology, 235(8), 2195– 2220. https://doi.org/10.1007/s00213-018-4950-4

30. Yamasaki, S., Aso, T., Miyata, J., Sugihara, G., Hazama, M., Nemoto, K., Yoshihara, Y., Matsumoto, Y., Okada, T., Togashi, K., Murai, T., Takahashi, H., & Suwa, T. (2020). Early and late effects of electroconvulsive therapy associated with different temporal lobe structures. Translational Psychiatry, 10(1), 344. https://doi.org/10.1038/s41398-020-01025-8

31. Fridgeirsson, E. A., Deng, Z.-D., Denys, D., van Waarde, J. A., & van Wingen, G. A. (2021). Electric field strength induced by electroconvulsive therapy is associated with clinical outcome. NeuroImage: Clinical, 30(30), 102581. https://doi.org/10.1016/j.nicl.2021.102581

32. Takamiya, A., Bouckaert, F., Laroy, M., Blommaert, J., Radwan, A., Khatoun, A., Deng, Z.-D., Mc Laughlin, M., Van Paesschen, W., De Winter, F.-L., Van den Stock, J., Sunaert, S., Sienaert, P., Vandenbulcke, M., & Emsell, L. (2021). Biophysical mechanisms of electroconvulsive therapy-induced volume expansion in the medial temporal lobe: A longitudinal in vivo human imaging study. Brain Stimulation, 14(4), 1038–1047. https://doi.org/10.1016/j.brs.2021.06.011

33. Sun, H., Jiang, R., Qi, S., Narr, K. L., Wade, B. S., Upston, J., Espinoza, R., Jones, T., Calhoun, V. D., Abbott, C. C., & Sui, J. (2020). Preliminary prediction of individual response to electroconvulsive therapy using whole-brain functional magnetic resonance imaging data. NeuroImage. Clinical, 26, 102080. https://doi.org/10.1016/j.nicl.2019.102080

34. Zuo, X.-N., & Xing, X.-X. (2014). Test-retest reliabilities of resting-state FMRI measurements in human brain functional connectomics: A systems neuroscience perspective. Neuroscience & Biobehavioral Reviews, 45(45), 100–118. https://doi.org/10.1016/j.neubiorev.2014.05.009

35. Conwell, K., von Reutern, B., Richter, N., Kukolja, J., Fink, G. R., & Onur, O. A. (2018). Test-retest variability of resting-state networks in healthy aging and prodromal Alzheimer’s disease. NeuroImage : Clinical, 19(19), 948–962. https://doi.org/10.1016/j.nicl.2018.06.016

36. Holiga, Š., Sambataro, F., Luzy, C., Greig, G., Sarkar, N., Renken, R. J., Marsman, J.-B. C., Schobel, S. A., Bertolino, A., & Dukart, J. (2018). Test-retest reliability of task-based and resting-state blood oxygen level dependence and cerebral blood flow measures. PLoS ONE, 13(11), e0206583. https://doi.org/10.1371/journal.pone.0206583

37. Cremers, H. R., Wager, T. D., & Yarkoni, T. (2017). The relation between statistical power and inference in fMRI. PloS one, 12(11), e0184923. https://doi.org/10.1371/journal.pone.0184923

38. Cléry-Melin, M., Jollant, F., & Gorwood, P. (2019). Reward systems and cognitions in Major Depressive Disorder. CNS Spectrums, 24(1), 64–77. doi:10.1017/S1092852918001335

39. Gabbay, V., Ely, B. A., Li, Q., Bangaru, S. D., Panzer, A. M., Alonso, C. M., Castellanos, F. X., & Milham, M. P. (2013). Striatum-based circuitry of adolescent depression and anhedonia. Journal of the American Academy of Child and Adolescent Psychiatry, 52(6), 628–41.e13. https://doi.org/10.1016/j.jaac.2013.04.003

40. Hahn, A., Haeusler, D., Kraus, C., Höflich, A. S., Kranz, G. S., Baldinger, P., Savli, M., Mitterhauser, M., Wadsak, W., Karanikas, G., Kasper, S., & Lanzenberger, R. (2014). Attenuated serotonin transporter association between dorsal raphe and ventral striatum in major depression. Human brain mapping, 35(8), 3857–3866. https://doi.org/10.1002/hbm.22442

41. Dandekar, M., Fenoy, A., Carvalho, A. et al. Deep brain stimulation for treatment-resistant depression: an integrative review of preclinical and clinical findings and translational implications. Mol Psychiatry 23, 1094–1112 (2018). https://doi.org/10.1038/mp.2018.2

42. Zhang, H., Li, L., Wu, M., Chen, Z., Hu, X., Chen, Y., … & Gong, Q. (2016). Brain gray matter alterations in first episodes of depression: a meta-analysis of whole-brain studies. Neuroscience & Biobehavioral Reviews, 60, 43–50.

43. Chen, M. H., Li, C. T., Lin, W. C., Hong, C. J., Tu, P. C., Bai, Y. M., Cheng, C. M., & Su, T. P. (2018). Persistent antidepressant effect of low-dose ketamine and activation in the supplementary motor area and anterior cingulate cortex in treatment-resistant depression: A randomized control study. Journal of affective disorders, 225, 709–714. https://doi.org/10.1016/j.jad.2017.09.008

44. Bennabi, D., Vandel, P., Papaxanthis, C., Pozzo, T., & Haffen, E. (2013). Psychomotor retardation in depression: a systematic review of diagnostic, pathophysiologic, and therapeutic implications. BioMed research international, 2013, 158746. https://doi.org/10.1155/2013/158746

45. Chen, C., Liu, Z., Zuo, J., Xi, C., Long, Y., Li, M. D., Ouyang, X., & Yang, J. (2021). Decreased Cortical Folding of the Fusiform Gyrus and Its Hypoconnectivity with Sensorimotor Areas in Major Depressive Disorder. Journal of affective disorders, 295, 657–664. https://doi.org/10.1016/j.jad.2021.08.148

46. Wang, J., Wei, Q., Bai, T., Zhou, X., Sun, H., Becker, B., Tian, Y., Wang, K., & Kendrick, K. (2017). Electroconvulsive therapy selectively enhanced feedforward connectivity from fusiform face area to amygdala in major depressive disorder. Social cognitive and affective neuroscience, 12(12), 1983–1992.

47. Hilland, E., Landrø, N. I., Kraft, B., Tamnes, C. K., Fried, E. I., Maglanoc, L. A., & Jonassen, R. (2019). Exploring the links between specific depression symptoms and brain structure: A network study. Psychiatry and clinical neurosciences, 74(3), 220–221.

48. Scangos, K. W., Weiner, R. D., Coffey, E. C., & Krystal, A. D. (2018). An Electrophysiological Biomarker That May Predict Treatment Response to ECT. The Journal of ECT, 35(2), 95–102. https://doi.org/10.1097/yct.0000000000000557

49. Hill, A. T., Hadas, I., Zomorrodi, R., Voineskos, D., Farzan, F., Fitzgerald, P. B., Blumberger, D. M., & Daskalakis, Z. J. (2020). Modulation of functional network properties in major depressive disorder following electroconvulsive therapy (ECT): a resting-state EEG analysis. Scientific Reports, 10(1). https://doi.org/10.1038/s41598-020-74103-y

50. Lanzenberger, R., Baldinger, P., Hahn, A., Ungersboeck, J., Mitterhauser, M., Winkler, D., Micskei, Z., Stein, P., Karanikas, G., Wadsak, W., Kasper, S., & Frey, R. (2012). Global decrease of serotonin-1A receptor binding after electroconvulsive therapy in major depression measured by PET. Molecular Psychiatry, 18(1), 93–100. https://doi.org/10.1038/mp.2012.93

51. Baldinger, P., Lotan, A., Frey, R., Kasper, S., Lerer, B., & Lanzenberger, R. (2014). Neurotransmitters and electroconvulsive therapy. The Journal of ECT, 30(2), 116– 121. https://doi.org/10.1097/YCT.0000000000000138

